# Upregulation of Activation Induced Cell Markers (AIM) among Severe COVID-19 patients in Bangladesh

**DOI:** 10.1101/2022.07.12.22276726

**Authors:** Taufiqur Rahman Bhuiyan, Hasan Al Banna, M Hasanul Kaisar, Polash Chandra Karmakar, Al Hakim, Afroza Akter, Tasnuva Ahmed, Imam Tauheed, Shaumik Islam, Mohammad Abul Hasnat, Mostafa Aziz Sumon, Asif Rashed, Shuvro Ghosh, John D Clemens, Sayera Banu, Tahmina Shirin, Daniela Weiskopf, Alessandro Sette, Fahima Chowdhury, Firdausi Qadri

**Affiliations:** International Centre for Diarrhoeal Disease Research Bangladesh (icddr,b), Dhaka, Bangladesh; Department of Genetic Engineering and Biotechnology, Faculty of Life and Earth Sciences, Jagannath University, Dhaka, Bangladesh; Kurmitola General Hospital, Dhaka, Bangladesh; Mugda Medical College & Hospital, Dhaka, Bangladesh; UCLA Fielding School of Public Health, Los Angeles, CA, USA; International Vaccine Institute, Seoul, South Korea; Institute of Epidemiology, Disease Control and Research, Dhaka, Bangladesh; Center for Infectious Disease and Vaccine Research, La Jolla Institute for Immunology (LJI), La Jolla, CA 92037, USA; Department of Medicine, Division of Infectious Diseases and Global Public Health, University of California, San Diego (UCSD), La Jolla, CA 92037, USA

**Keywords:** COVID-19, SARS-CoV-2, Bangladesh, Activation Induced Marker, CD4^+^ T cells, MAIT cells

## Abstract

COVID-19 caused by SARS-CoV-2 can develop the disease with different degree of clinical severity including fatality. In addition to antibody responses the antigen specific T cells may play a critical role in defining this protective immune response against this virus. As a part of a longitudinal cohort study in Bangladesh to investigate B and T cell specific immune responses, we sought to evaluate the activation induced cell marker (AIM) and the status of different immune cell subsets during infection. A total of 115 participants were analyzed in this study which included participants with asymptomatic, mild, moderate and severe clinical symptoms. In addition, healthy controls (19 in each group) were analysed. Specimens from participants collected during the pre-pandemic period were also analyzed (n=10). Follow-up visits were conducted on day 7, 14, and 28 for all the cases since the enrollment (day 1). In this study 10 participants among the moderate and severe cases expired during the course of follow up. We observed a decrease in mucosa associated invariant T (MAIT) cell frequency on the initial days (day 1 and day 7) in comparison to later days of the COVID-19 infection. However, natural killer (NK) cells were found to be elevated in symptomatic patients just after the onset of disease compared to both asymptomatic patients and healthy individuals. Moreover, we found AIM^+^ (both OX40^+^ CD137^+^ and OX40^+^ CD40L^+^) CD4^+^ T cells to show significant increase in moderate and severe COVID-19 patients in response to SARS-CoV-2 peptides (specially spike peptide) compared to prepandemic controls, who are unexposed to SARS-CoV-2. Notably, we did not observe any significant difference in the CD8^+^ AIM markers (CD137^+^ CD69^+^), which indicates the exhaustion of CD8^+^ T cells during COVID-19 infection. These findings suggest that the patients who recovered from moderate and severe COVID-19 were able to mount a strong CD4^+^ T cell response against shared viral determinants that ultimately induced the T cells to mount further immune responses to SARS-CoV-2.

## INTRODUCTION

The coronavirus disease 2019 (COVID-19) caused by the novel coronavirus SARS-CoV-2 has created an unprecedented global pandemic causing over 364 million confirmed cases and over 5.6 million deaths as of January 28, 2022 (1). In spite of a number of available vaccines against the COVID-19 the spread of the disease is still not controlled everywhere. This disease shows several stages of severity, different systemic nature from other respiratory diseases, and unpredictable outcome with possible comorbidities including cardiovascular disease, obesity, diabetes, lung diseases, kidney diseases and cancer. These have made the management of COVID-19 patients challenging. Therefore, an understanding of human T-cell response to SARS-CoV-2 is imperative to quantify the antigen specific CD4^+^ and CD8^+^ T cells which will provide insights into immunity and pathogenesis of SARS-CoV-2 infection as well as vaccine design and evaluation of candidate vaccines (2). There are a number of assays conventionally used for measuring the quantity and quality of antigen-specific T cells in humans, either at the single cell level by flow cytometry or enzyme-linked immunospot (ELISpot) and intracellular cytokine staining (ICS) assays or at the population level by enzyme-linked immunosorbent assay (ELISA) (3-5). Several research groups have overcome the limitations by antigen specific T cells assays based on the upregulation of T-cell receptor (TCR)-stimulated surface markers called activation-induced markers (AIM), which can effectively determine the overall antigen-specific T cell response (6). Several studies have been successfully used the AIM assays to detect the virus-specific, vaccine-specific, or tuberculosis-specific CD4+ T cells (7-9).

Pre-existing immunity acquired from ‘common cold’ Human Coronaviruses (HCoVs) could have extensive consequence in the outcome of the pandemic. Recent reports reveal the existence of cross-reactive CD4^+^ T cells in ∼20-50% of the individuals who have not been exposed to SARS-CoV-2 (10, 11). In fact, less is known in relation to cross-reactive immune memory to SARS-CoV-2 CD4^+^ T cells, antibody analyses and more studies are required to studied as no firm data are available yet (12, 13). However, a concern has been arisen because a decline of antibodies have been observed within first few months after recovery from SARS-CoV-2 infection (14, 15). Moreover, identifying the particular immune cells that are being activated by SARS-CoV-2, may have substantial impact on evaluating the immune response to COVID-19 vaccines. Therefore, to understand the attributes of adaptive immunity, in this study, we have examined the activation induced markers responses and stability of immune memory in unexposed donors and patients recovered from COVID-19 infection for observing COVID-19 specific immune responses.

## METHODS AND MATERIALS

### Participants and Study Design

Participants of this study were classified in three groups-unexposed individuals, healthy control and COVID-19 patients (19). COVID-19 infected patients were followed up for 1 year at day 1, day 7, day 14, day 28, day 90, day 180, day 270 and day 360 after becoming RT-PCR positive. Patients were enrolled before starting COVID vaccination in Bangladesh. For unexposed individuals, we used cryopreserved Peripheral Blood Mononuclear Cells (PBMC) from healthy participants collected before the pandemic era. The healthy controls and COVID-19 participants of this study are part of an ongoing longitudinal cohort study, which has been conducted in Dhaka, Bangladesh since November, 2020. COVID-19 patients were enrolled from two COVID-19 hospitals, Mugda Medical College and Hospital and Kurmitola General Hospital and the non-hospitalized patients were enrolled from the community in and around the Dhaka city (19). Nasopharyngeal swab samples were collected from the patients to diagnose the SARS-CoV-2 by reverse transcription polymerase chain reaction (RT-PCR). Participants who had never been diagnosed for COVID-19, had no clinical signs and symptoms for at least 2 weeks and were tested negative by RT-PCR for SARS-CoV-2 before the enrollment, were considered as healthy controls. We have enrolled healthy participants during the same time when we have enrolled COVID-19 patients by matching age and gender. Among the total 115 participants of this study 86 were COVID-19 cases of four types of different disease severity (n=19) in each group, along with 10 who had expired due to COVID-19, 19 healthy individuals and 10 unexposed or pre-pandemic participants (collected before October 2019). A fraction of COVID-19 patients from each category was used for AIM assay on multiple day points which includes asymptomatic (n=5), mild (n=6, every fourth), moderate (n=6, every fourth), severe (n=15, every alternate), and expired (n=10, all) patients. The AIM assay was also conducted in 10 unexposed and 9 healthy participants. The existing WHO standards (16) were used to classify the patients’ disease severity, which was determined from their hospital admission records or clinical status at the time of study enrolment. Asymptomatic cases were classified as those who tested positive for RT-PCR but had no signs or symptoms of illness. We randomly tested RT-PCR in the participants in the community and who were positive but having no COVID symptoms were enrolled as asymptomatic participants. The participants had a mean age of 45.86 years (n=115, range 18 to 76 years). Among all the participants, 70 were male and 45 were female. A detail of all demographic information (age, sex, blood group) for participants’ of the AIM assay (n=61) were given in the **Supplementary Table 1**. This study was approved by the Institutional Review Board (IRB) of icddr,b and the Directorate General of Health Services (DGHS), Bangladesh. Written consent was collected from the study participants before enrollment.

### Isolation of Peripheral Blood Mononuclear Cells (PBMCs)

Blood samples were collected in Sodium-EDTA tube and isolation of peripheral blood mononuclear cells (PBMCs) was done by density gradient method using Ficoll-Paque PLUS (Cytiva, USA). Before isolating cells, plasma was separated using centrifugation. After separatingt plasma, the whole blood was diluted as 1:1 ratio using R-10 media (89% RPMI 1640 (Gibco), 10% Fetal Bovine Serum (FBS; Gibco), 1% Penicillin-Streptomycin (Gibco)). Isolated PBMCs were resuspended to a concentration of 1 × 10^6^ cells/mL in RPMI complete medium (87% RPMI 1640, 10% FBS, 1% Penicillin-Streptomycin, 1% Sodium-Pyruvate (Gibco), 1% L-Glutamine (Gibco)). Fraction of the cells were used as fresh for flow cytometry and rest of the cells were cryopreserved using the cryoprotectant (90% FBS, 10% DMSO). These cryopreserved cells were later used in different immune assays, including the AIM assay.

### Flow Cytometry and T Cell Phenotyping

All immunophenotyping experiments were performed on FACSAria Fusion instruments (BD Bioscience, San Jose, CA). Freshly separated PBMCs were stained with fluorochrome-tagged antibodies which are listed in the **Supplementary Table 2**. After staining the cells were washed with PBS with 2% FBS (FACS buffer) and antibody tagged cells were fixed using Cytofix (BD Biosciences, Cat# 554655). Then the data were acquired using the FACSDiva software program the next day. During analyses live singlet lymphocytes were gated to quantitate different types of T cells (See representative gating in **Supplementary Figure1**).

Helper T cell were gated as CD19^-ve^ CD3^+ve^ CD4^+ve^ cells and cytotoxic T cells were gated as CD19^-ve^ CD3^+ve^ CD8^+ve^. Follicular helper T cells are CXCR5^+ve^ cells gated on helper T cells. Fluorochrome-tagged antibodies that were used to stain Mucosal-associated Invariant T (MAIT) cells are listed in the **Supplementary Table 3**. MAIT cells were gated on live singlet as CD3^+ve^ CD8^+ve^ TCR Vα7.2^+ve^ CD161^+ve^ (See representative gating in **Supplementary Figure2**). Again, antibodies used for Natural Killer (NK) cell staining are listed in the **Supplementary Table 4**. During NK cell analysis CD19 and CD14 were dumped using a single channel and then the NK cells were gated as CD3^-ve^ CD16^+ve^ CD56^+ve^. Lastly, the central memory (Tcm) and the effector memory (Tem) T cells were gated as CD45RO^+ve^ CD27^+ve^ and CD45RO^+ve^ CD27^-ve^ respectively on either CD4 or CD8 cell. All the phenotyping results are expressed as percentage of parent population. We have analyzed the acquired data with FlowJo software (version 10.6.1, TreeStar Inc., USA).

### Activation Induced Marker (AIM) Assay

To identify and quantitate the SARS-CoV-2−specific CD4^+^ as well as CD8^+^ T cell responses, we performed T cell receptor dependent (TCR) dependent AIM assays (2) using cryopreserved PBMCs. We stimulated PBMC of COVID-19 patients and other control samples with four different SARS-CoV-2 specific peptide megapools (MPs). SARS-CoV-2 specific CD4 peptide MPs were divided into two groups: spike protein as spike MP and all other polyproteins as non-spike MPs. Again, the antigen-specific analyses of CD8 T cells were performed using the class I peptide megapool prepared from the whole virus proteome and consist of 628 peptides. The megapool splited into two parts consisting of 314 peptide each, CD8-A MP and CD8-B MP (17). Cryopreserved cells were thawed and were rested overnight in 37°C incubator supplemented with 5% CO_2_. Next day, they were distributed into 96-well U bottom plate in a total of 0.3×10^6^ PBMCs per well and different peptide MPs were added at 1 mg/mL concentration. An equimolar quantity of DMSO (vehicle) and phytohaemagglutinin (PHA) were used as negative and positive controls, respectively. Moreover, we used Cytomegalovirus derived peptides, like-CMV CD4 MP and CMV CD8 MP as positive control to compare the stimulations with the SARS-CoV-2 MPs. The PBMCs were stimulated for 24 hours in a 37°C incubator supplemented with 5% CO_2_. The stimulated cells were stained for 1 hour at 4 °C in the dark with a cocktail of antibody panel (**Supplementary Table 5**) targeting the AIM^+^ T cells. Following the surface staining, cells were washed with PBS with 2% FBS and fixed using Cytofix (BD Biosciences). Later data acquisition was carried out in a FACSAria Fusion cytometer through FACSDiva software. Data analyses was done using FlowJo 10.6.1. During analysis live singlet lymphocytes were gated and CD14^+ve^ and CD16^+ve^ cells were discarded by a dump channel. Then both OX40^+ve^CD40L^+ve^ and OX40^+ve^CD137^+ve^ cells were gated on CD4^+ve^ cells, while CD137^+ve^CD69^+ve^ cells were gated on CD8^+ve^ cells (see representative gating in **Supplementary Figure 3**). We measured Stimulation Index (SI) for each participant for every type of stimulation to quantify the extent of stimulation. SI has been defined by dividing the percentage of megapool stimulated AIM^+^ cells with the percentage of DMSO stimulated AIM^+^ cells. Stimulation due to SARS-CoV-2 CD8 MP was determined using the combined SI data of CD8-A and CD8-B MP.

### Statistical Analysis

All statistical analysis was performed using the GraphPad Prism 6.0. Flow cytometry figures were generated using FlowJo software and other plots were generated using GraphPad Prism. To compare DMSO versus spike, non-spike, CMV, or PHA in both Unexposed and COVID-19 cases we used Wilcoxon matched-pairs signed rank test because the data was paired and nonparametric. To compare unexposed versus COVID-19 cases we performed two-tail Mann-Whitney test because the data was unpaired and nonparametric. We also performed Mann-Whitney test while comparing immune cell frequency in symptomatic and asymptomatic patients. Again, while comparing different day points of the same category of disease condition we utilized paired t test as the data was parametric. Moreover, we conducted a one-way ANOVA to compare immune cell frequency of multiple groups with a single group (healthy control) followed by Dunnett’s Multiple Comparison Tests to see the differences compared to control. All tests are specified in the corresponding figure legends. All the statistical analyses were done considering 95% confidence level.

## RESULTS

### Frequency of Helper, Cytotoxic, and Follicular Helper T cells with Disease Severity and Time Points

We conducted phenotypic analyses of fresh PBMCs for different subsets of T cells We did not observe any statistically significant difference in frequency for helper, cytotoxic, and follicular T cells among different disease categories in different day points from disease onset (**Figure 1 A-C**).

**Figure 1:**
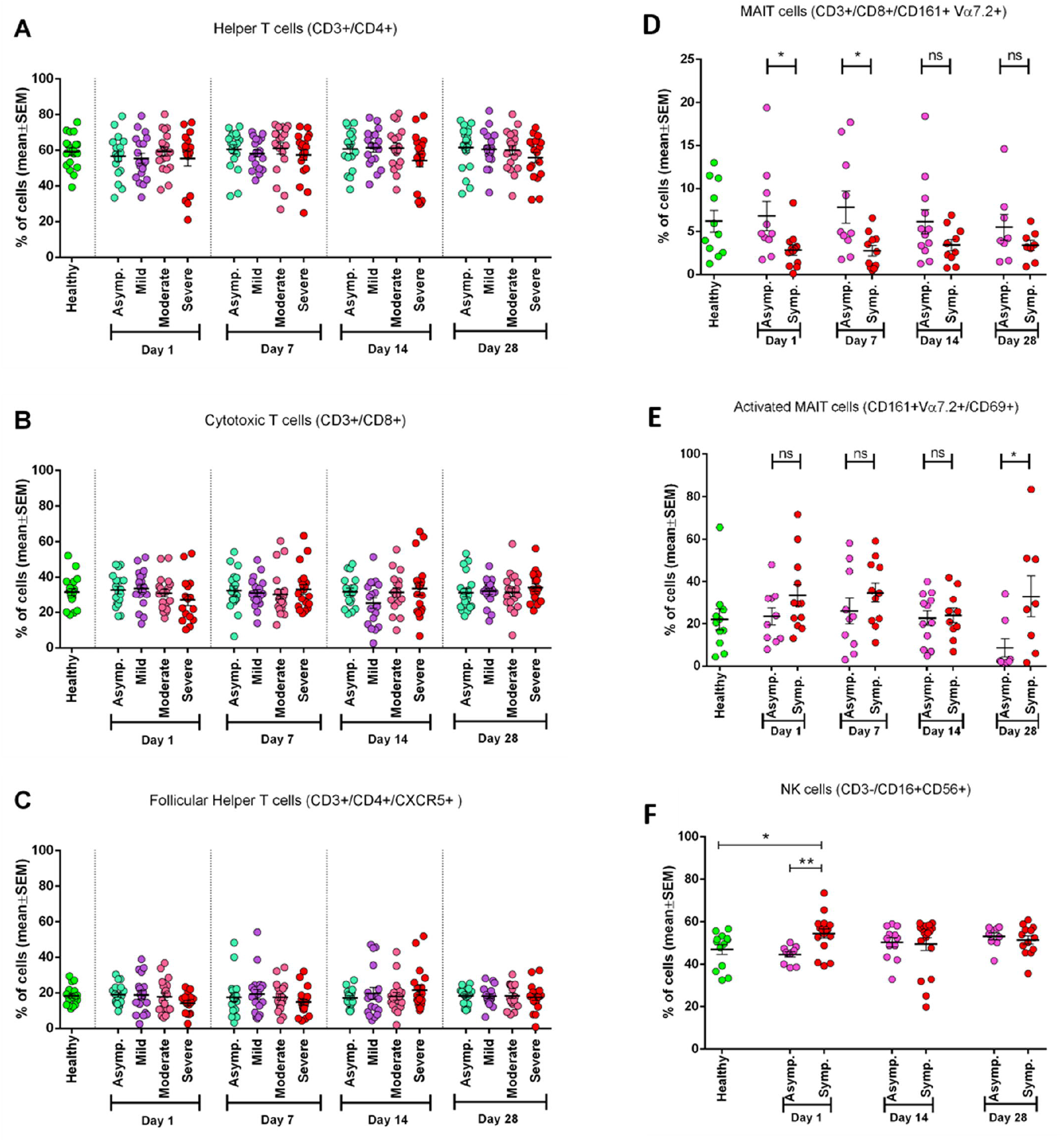
Frequency of different immune cell types in different category of COVID-19 patients (Asymptomatic, Mild, Moderate, and Severe) in different day points from the onset of infection and compared with the Healthy controls. (A) Percentage of Helper T cells in CD3^+^ cells, (B) percentage of Cytotoxic T cells in CD3^+^ cells, (C) percentage of Follicular Helper T cells in CD4^+^ cells. In plots (D-E) symptomatic and asymptomatic COVID-19 patients are compared for-(D) MAIT cells as a percentage of CD8^+^ cells, (E) CD69^+^ activated MAIT cells. (F) Percentage of Natural Killer (NK) cells. Bars represent mean value with Standard Error of Mean. Statistical comparisons were done (D-F) using two-tail Mann-Whitney test. *p <0.05; **p <0.01; ***p < 0.001; ns: non-significant.

### MAIT Cell Frequency in Symptomatic Patients During the Initial Days of Infection

We have found that MAIT cell frequency tend to decrease significantly (p= 0.0111) in the initial days of infection (both in Day 1 and Day 7). In later days (Day 14 and Day 28) this difference become statistically insignificant in symptomatic patients compared to both asymptomatic patients and healthy participants. (**Figure 1D**). We further investigated MAIT cells by including an activation marker (CD69) and observed a higher expression of the marker remains in symptomatic patients throughout the infection compared to asymptomatic patients. Even the difference gets statistically significant (p <0.05) after one month (Day 28) (**Figure 1E**).

### Natural Killer (NK) Cells in Symptomatic Patients

During the early days of infection, we observed significantly higher frequency of NK cell in symptomatic patients (P= 0.0014) compared to asymptomatic patients. We also found that NK cell activity was significantly higher in symptomatic patients compared to healthy controls, as expected. However, this difference did not remain significant in the later day points of infection (day 14 and 28) (**Figure 1F**).

### Frequency of CD4+ and CD8+ Central and Effector Memory T Cells

To see the status of Central Memory T cell (Tcm) and Effector Memory T cell (Tem), we measured their frequency in both CD4^+^ and CD8^+^ T cells (**Figure 2)**. We found that the frequency of CD4^+^ Tcm cells significantly (p= 0.0119) differs from the control group, but CD4^+^ Tem cells do not (**Figure 2A, B)**. On the other hand, we have got the statistically significant (p= 0.0055) difference for CD8^+^ Tem cells, and not for CD8^+^ Tcm cells, when compared with the healthy control group (**Figure 2C, D**).

**Figure 2:**
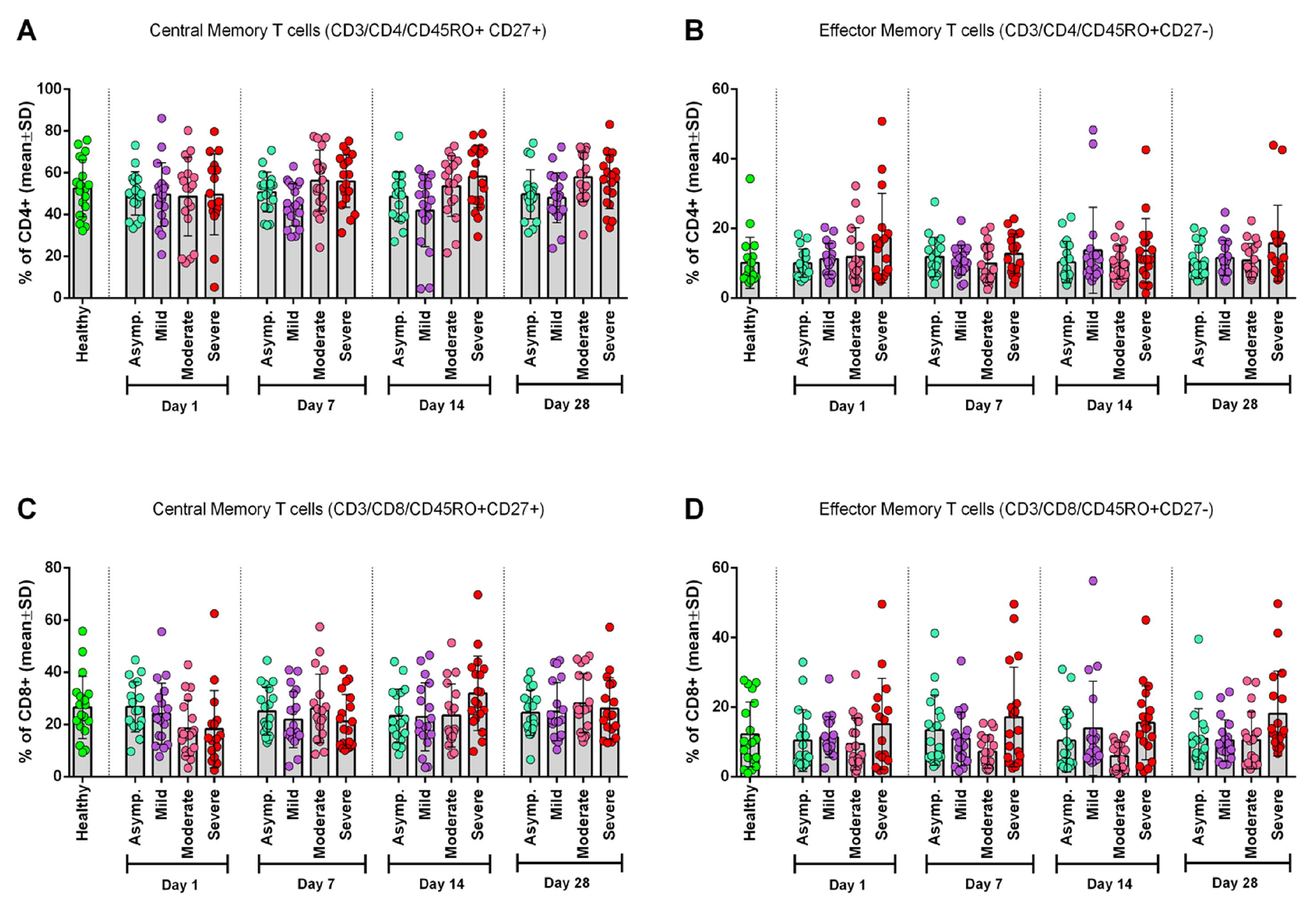
Frequency of Memory T cell subtypes in different category of COVID-19 patients (Asymptomatic, Mild, Moderate, and Severe) in different day points from the onset of infection and compared with the Healthy controls. (A) CD4^+^ Central Memory T cells, (B) CD4^+^ Effector Memory T cells, (C) CD8^+^ Central Memory T cells, and (D) CD8^+^ Effector Memory T cells. One-way ANOVA was performed for each of the plots to compare differences of each column with corresponding Healthy control data.

### Cell Recovery Tends to Decrease After Thawing for Severe Patients

While we thawed cryopreserved PBMCs for AIM assays, we observed very low count of PBMCs at their day 1 in case of Severe COVID-19 patients compared to healthy and other categories. This trend remains the same after overnight resting at 37°C in the CO_2_ incubator. Therefore, we plotted the values and found a significant decrease in recovery of the PBMCs in severe patient’s compered to healthy control (**Supplementary Figure 4A**). Recovery after thawing from day 1 PBMCs of unexposed, healthy, asymptomatic and dead participants showed no significant differences. This scenario led us to analyze the number of PBMCs per mL of blood of these samples before cryopreservation and we again found significantly less number of PBMCs from day 1 samples of severe cases compared to healthy control (**Supplementary Figure 4C**). PBMCs of day 28 from the different patient groups did not show any remarkable difference in case of the percentages of recovery and number of PBMCs per mL of blood (**Supplementary Figure 4B, 4D**).

### SARS-CoV-2-Specific CD4^+^ T Cell Responses in Severe COVID-19 Patients

To explore the T cells primed for anti-viral immune response, we have evaluated the SARS-CoV-2 specific CD4^+^ T cells in the TCR dependent AIM assay in COVID-19 patients of different disease severity. We have found that AIM^+^ (OX40^+^ CD137^+^) CD4^+^ T cells showed significant increase over the DMSO control in response to both peptide MP spanning the spike domain (spike) and the MP covering the remainder of the SARS-CoV-2 genome (non-spike) (**Figure 3A, B**). When compared between the unexposed response to OX40^+^ CD137^+^ with recovered severe patients (Day 28), we found statistically significant differences for spike (p= 0.002), but not for non-spike MPs (**Figure 4B**). Again, both SARS-CoV-2 spike and non-spike reactive AIM^+^ (OX40^+^ CD40L^+^) CD4^+^ T cells have also shown significant increase over DMSO control (**Supplementary Figure 5**). Similarly, CMV MP and PHA was also used as a positive control here. Notably here SI was also found to be significantly higher for both spike (p=0.0007) and non-spike (p=0.0054) MPs in response to this (OX40^+^ CD40L^+^) AIM panel in severe COVID-19 patients compared to the unexposed participants (**Figure 4A**). In case of both AIM panels, the unexposed donors consistently responded to the CMV peptide MP and the PHA superantigen significantly over the DMSO control (**Supplementary Figure 5A, B**). So, according to our data, severe COVID-19 patients consistently a substantial CD4^+^ T cell response against SARS-CoV-2 after one month from the onset of the infection.

**Figure 3:**
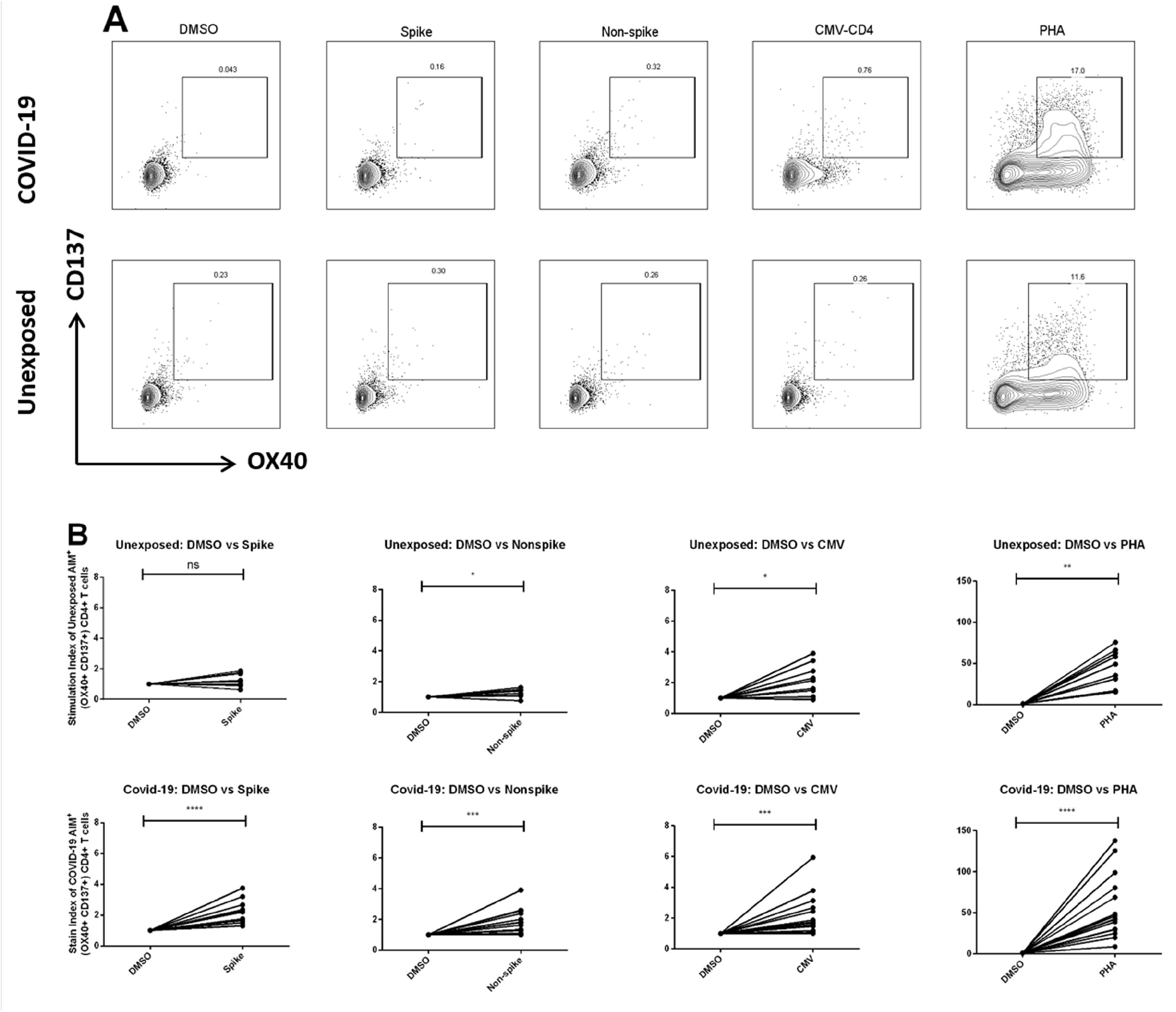
(A) Presentative plot for Fluorescence-activated cell sorting (FACS) gating for AIM^+^ (OX40^+^ CD137^+^) cells gated on CD4^+^ T cell; (B) AIM^+^ CD4^+^ T cell reactivity in COVID-19 cases between the negative control (DMSO) and different antigen-specific stimulations (Spike, Non-spike MP, CMV, PHA). Wilcoxon matched-pairs signed rank test was performed to compare between groups. *p <0.05; **p <0.01; ***p < 0.001; ns: non-significant.

**Figure 4:**
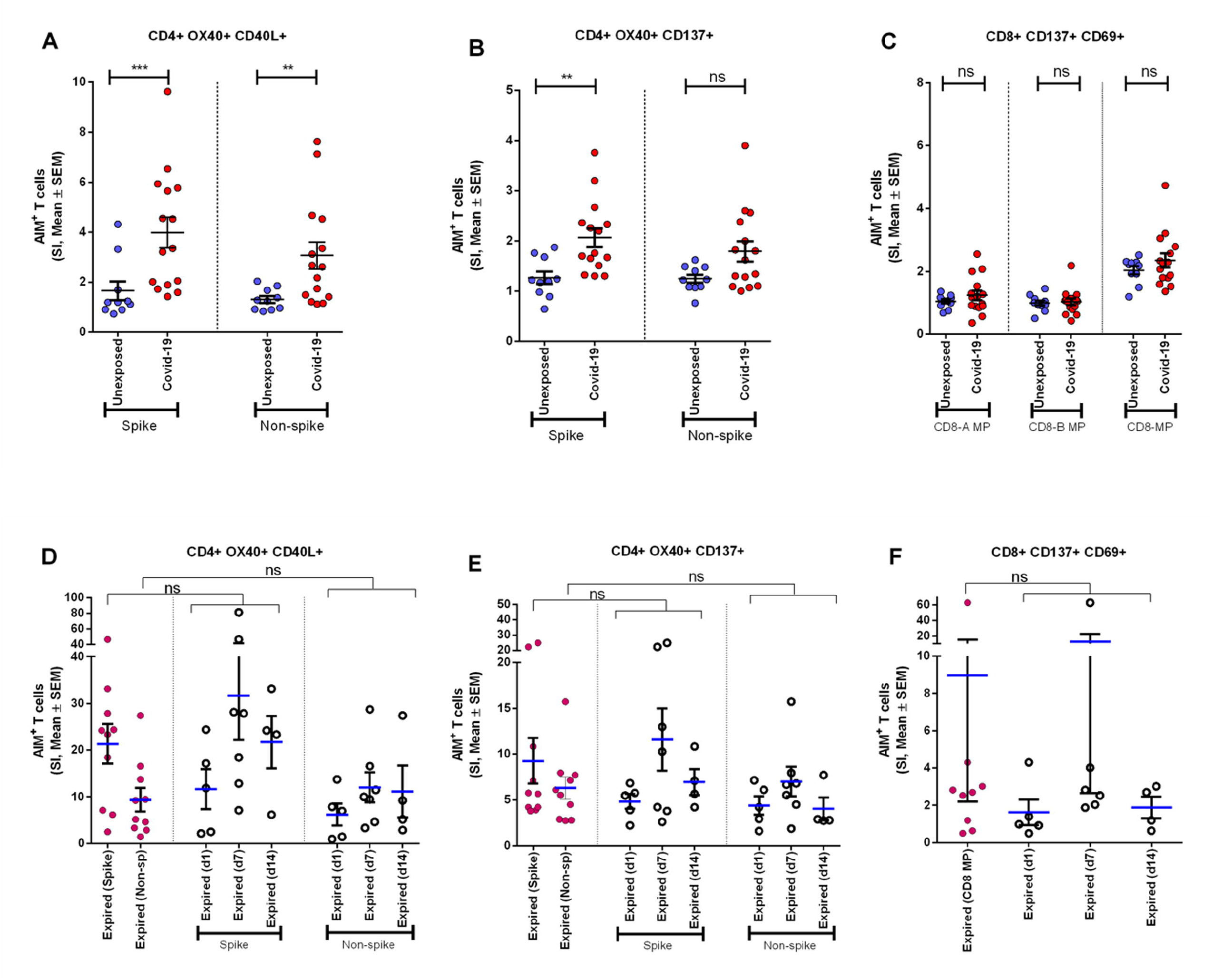
Antigen specific response to different CD4^+^ and CD^+^ AIM markers. (A-C) AIM expression in Unexposed & Covid-19 participants. Stimulation Index (SI) quantitation of the AIM^+^ T cells after stimulation with CD4-nCOV-Spike, Non-spike (CD4-nCOV-all MP), and the class I CD8 peptide MPs (CD8-A and CD8-B). Covid-19 patient samples are collected after one month (day 28) after infection. For Unexposed n=10, and for Covid-19 n=15. (D-F) AIM expression in “Expired” participants. The left column (purple) shows SI of expired participants (n=10) from samples collected immediate visit before their death. Ordinary one way ANOVA (with Tukey’s multiple comparison test) was done to compare the groups. Figure shows the mean SI with error bars representing standard errors of the means (mean ± SEM). Statistical comparisons were performed by two-tail Mann-Whitney test. ns: non-significant, *p<0.05, **p<0.01, ***p<0.001

### SARS-CoV-2-Specific CD8^+^ T Cell Responses in Severe COVID-19 Patients

To measure the SARS-CoV-2 specific CD8^+^ T cells in unexposed and severe COVID-19 patients, we used CD8-A and CD8-B peptide MPs where whole virus proteome was split between these group of MPs. Though both unexposed and COVID-19 patients responded consistently to CMV and PHA, they did not respond to CD8-A and CD8-B MPs substantially (**Supplementary Figure 6A, B**). When we combined the SI for CD8-A and CD8-B MPs we got somewhat higher AIM^+^ (CD137^+^ CD69^+^) CD8^+^ T cell response, but that was not statistically significant (**Figure 4C**).

In essence, when comparing the recovered severe COVID-19 patients to the unexposed participants, antigen-specific T cell studies reveal a predominant role of CD4^+^ T cells over CD8^+^ T cells.

### SARS-CoV-2-Specific CD4^+^ & CD8^+^ T Cell Response in Patients who had Expired

We have compared AIM assay markers in COVID-19 patients who expired at different days after onset of symptoms. Very high levels of CD4^+^ AIM makers (OX40^+^137^+^, OX40^+^CD40L^+^) were observed at day 7 and day 14 after onset of symptoms to spike and non-spike proteins. Similarly, we observed CD8+ AIM markers (CD137+CD69+) was also upregulated at day 7 after onset of symptoms (**Figure 4D,E,F**). In addition, we used one way ANOVA to see the differences in response to Spike and Non-spike MPs among respective groups of day points. Mean of SI given by samples taken on the last visit before their death for all 10 expired patients showed no significant differences with the responses found in individual day points. This result indicates that deceased patients generally had a very high antigen specific T cell response before death and looked similar when compared combinedly as well as individually in different day points since infection.

### AIM^+^ T Cell Response to Spike Generates Over Time

To investigate the periodic scenario of the COVID-19 disease condition from every category, we performed the AIM assay for all category as well as for healthy controls in presence of both spike and non-spike MPs. When we compared the SI against spike MPs by paired analysis between Day 1 and Day 28 of infection, we found that every category responded significantly for AIM^+^ (OX40^+^ CD40L^+^) CD4^+^ T cells (**Figure 5**). This conveys the message that the T cell response augmented due to the COVID-19 infection over time. Though the AIM^+^ T cell response looks somewhat elevated in every case for Day 28, it was not statistically significant for neither non-spike MPs nor for another AIM panel (OX40^+^ CD137^+^).

**Figure 5:**
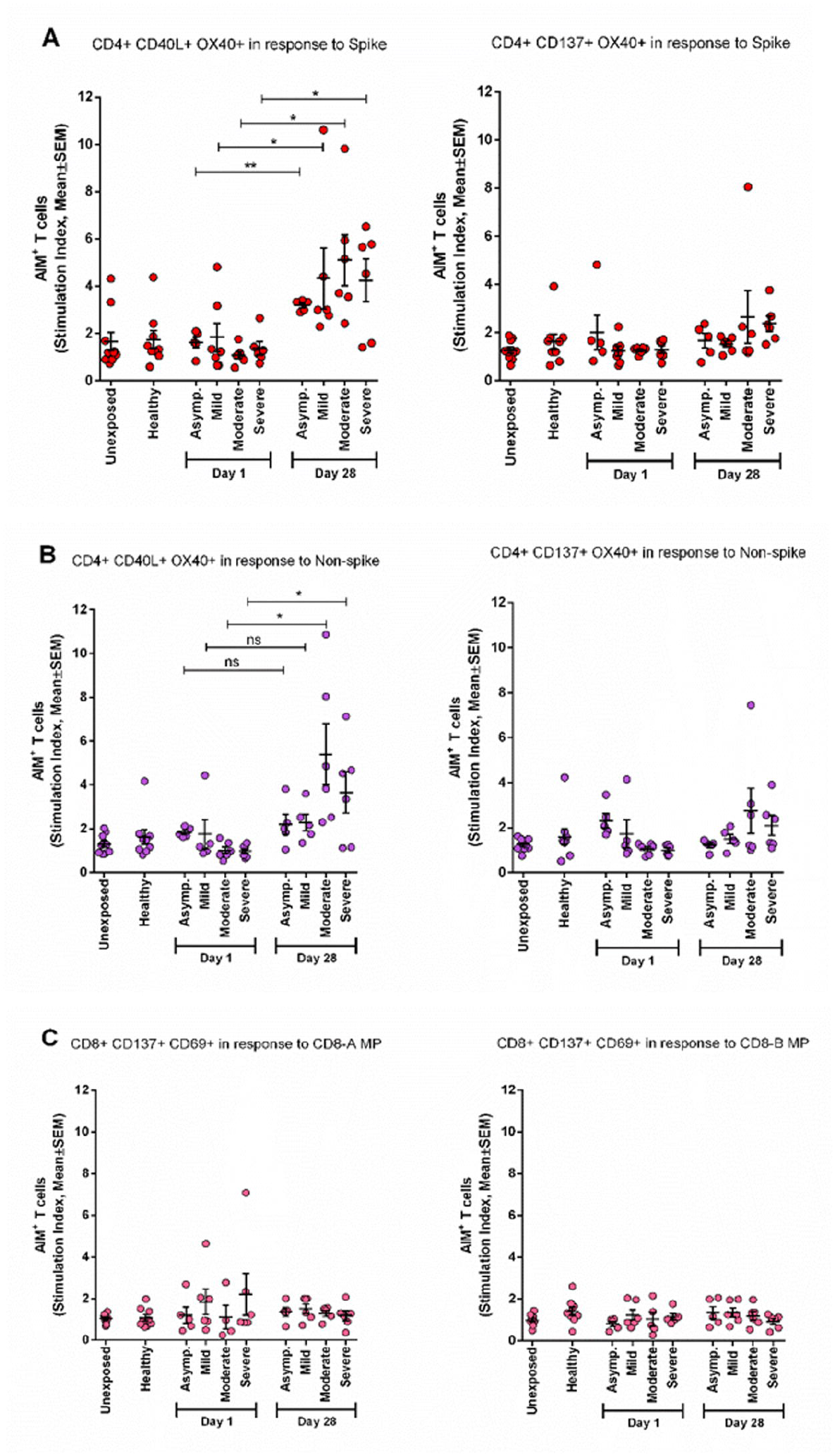
AIM expression in different category of COVID-19 patients immediately after infection (Day1) and after one month of infection (Day 28). (A) Stimulation of AIM+ (OX40^+^ CD40L^+^ and OX40^+^ CD137^+^) CD4^+^ T cells in response to Spike antigen; (B) Stimulation of AIM^+^ (OX40^+^ CD40L^+^ and OX40^+^ CD137^+^) CD4+ T cells in response to Non-spike megapool peptides; (C) Stimulation of AIM^+^ (CD137^+^ CD69^+^) CD8^+^ T cells in response to CD8-A and CD8-B megapool peptides. Paired t test is done to compare statistically groups. ns: non-significant, *p<0.05, **p<0.01, ***p<0.001

### Activation of Follicular Helper T cells (Tfh) Due to COVID-19

To determine the role of follicular helper T cell (Tfh) in recovered severe COVID-19, we performed the same stimulation experiment utilizing both spike and non-spike MPs and analyzed the cultured cells by flow cytometry using two different activation markers. We found that frequency of CXCR5^+^ CD40L^+^ cells tend to increase significantly (P < 0.05 for both spike and non-spike), while CXCR5^+^ PD1^+^ helper T cells tend to decrease, though not significantly, after one month of infection (**Figure 6**). This data suggests possibility of the role of Tfh cells during our immune system’s fight against SARS-CoV-2.

**Figure 6:**
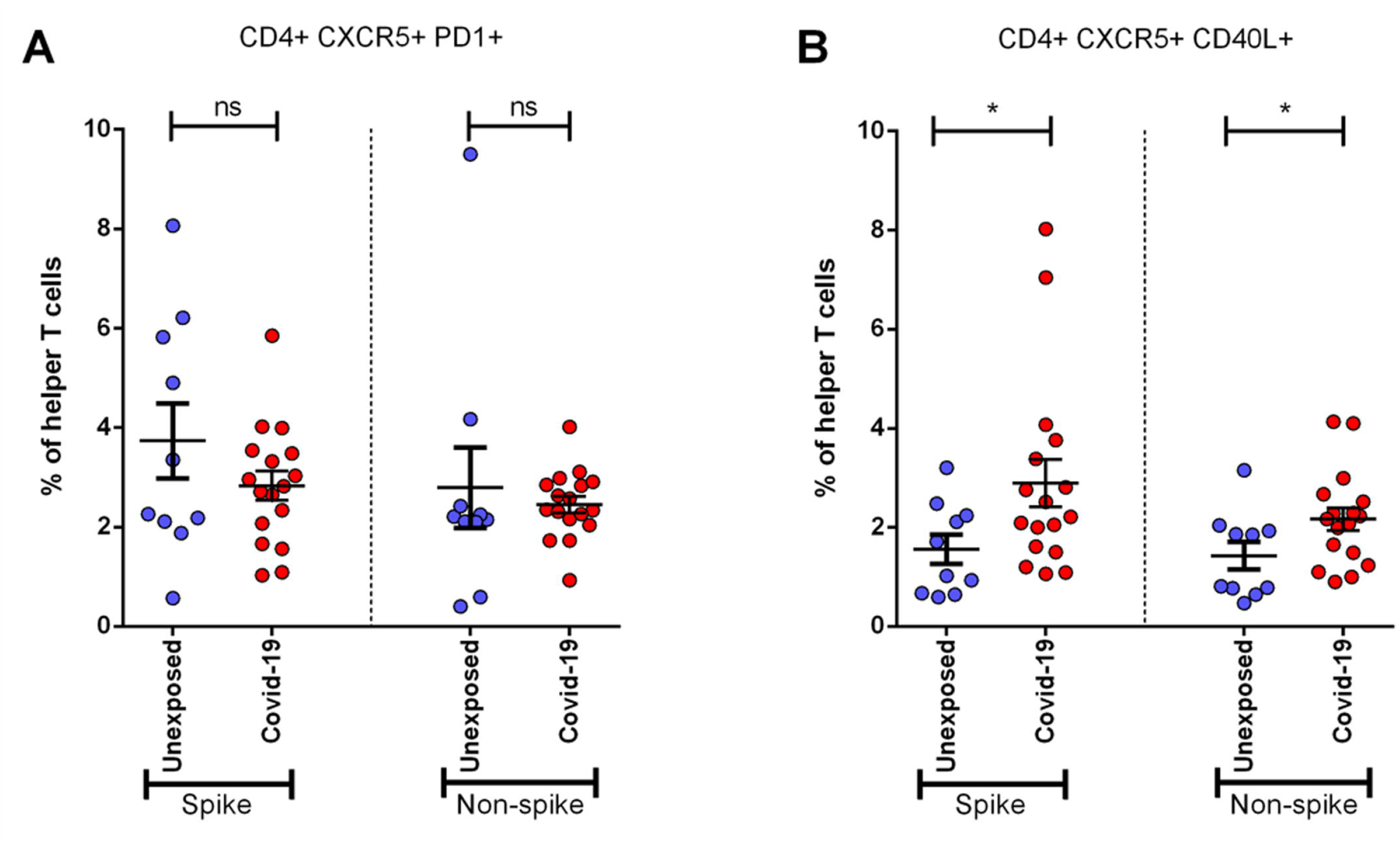
Activation of Follicular Helper T cells (Tfh) in Unexposed individual & severe COVID-19 patients. Samples were collected after one month (Day 28) of infection and response were observed for stimulation by both Spike and Non-spike antigens. (A) Frequency of CXCR5+ PD1^+^ cells in helper T cells; (B) Frequency of CXCR5^+^ CD40L^+^ cells in helper T cells. Statistical comparisons were done using two-tail Mann-Whitney test. *p<0.05; **p<0.01; ***p < 0.001; ns: non-significant.

## DISCUSSION

Understanding of adaptive immunity to COVID-19 has increased but remains limited and unclear especially in acute and convalescence COVID-19 patients. As the study was exploratory, the antigen-specific antibody and T cell data suggest the following conditions: (i) AIMs may limit COVID-19 severity; (ii) with prominent roles SARS-CoV-2-specific helper T cells are associated with less COVID-19 severity (18); and (iii) aging and lack of naive T cells number possibly linked to the unsuccessful synchronized AIM responses that result increased vulnerability to severe COVID-19. These findings have significant involvement both to understand immunity and pathology to novel coronavirus, as well as for designing an effective COVID-19 vaccine.

In our earlier reports, we have presented clinical, genomic and humoral antibody responses to Covid-19 (19-21) The is the first longitudinal study in Bangladesh that has been carried out to evaluate the adaptive T cell immune responses in patients with differing levels of severity of illness after SARS-CoV-2 infection. Bangladesh is similar to other countries globally, regarding age, gender and comorbid conditions (22, 23). However, the death rate due to COVID-19 is low in Bangladesh as the demographic characteristics of our population is different in compared to other countries, and elderly population (>50 years) is only 10-15% (24). Meanwhile, we would also like to observe the adaptive T cell responses in these different categories of severe patients and have compared the responses with peripheral blood mononuclear cells collected from a pre pandemic time point as health controls.

This study demonstrates the presence of robust antigen specific CD4+ T cell responses specific for SARS-CoV-2 in the COVID-19 recovered participants PBMCs. Higher viral loads may develop stronger SARS-CoV-2 specific T cell responses who had severe disease and may reflect a poor early T cell response that might be inadequate to clear/control the virus along with other factors including less innate immunity. Alternatively, it is possible that the T cell response was itself harmful and contributed to severity. Consistent with recent reports Alternatively, there is a probability that T cell response was itself harmful which might contribute severity. Consistent with current findings (2), (25), high frequency of helper T cell responses specific to spike protein was observed in recovered COVID-19 patients. This is much alike to influenza infection, where surface hemagglutinin of the virus elicited helper T cell responses, whereas the greater part of cytotoxic T cell responses found specific to viral internal proteins (26). CD8^+^ T cells recognize and kill host cells already infected by the virus and according to our results, they got exhausted after 1 month of infections which correspond to earlier study (27). Understanding the roles, timing and strength of different subsets of T cells in protection of SARS-CoV-2 is crucial for the prevention and treatment of COVID-19 infection. By the time, several vaccines are available and WHO approved globally and different countries including Bangladesh are administrating vaccine in the population and trying to increase the coverage rate as much as possible. The current COVID-19 infection is more challenging than prophylaxis. The data presented here suggest that SARS-CoV-2-specific CD4^+^ T cells has association in resolving acute COVID-19 infection. These findings suggest that introduction of vaccine can elicit both helper and cytotoxic T cells specific to SARS-CoV-2, along with protective neutralizing antibodies, and thus may generate immunity to provide adaptive antiviral immune response in SARS-CoV-2 infection. However, further understanding is required to determine adaptive T cell immune responses in current vaccinees simultaneously and correlate the responses with natural infection. By studying antigen-specific CD4 T cells, we can monitor the development of crucial immunological responses. As the helper T cell responses are significantly diverse, and thus detection of antigen-specific helper T cells with the production of one or more cytokines is likely to remarkably miscalculate the amount of the total antigen-specific response (28, 29).

In the current longitudinal study, we reported that moderate or severe disease patients had significantly higher antibody responses to SARS-CoV-2 RBD in compared to mild or asymptomatic infection (19). In further analyses, kinetics of IgG antibody has shown persistence until day 270 in moderate and severe patients and in contrast mild and asymptomatic participants antibody dropped from day 180 after onset of COVID-19 diagnosis. The IgM antibody responses showed transient immune responses and dropped after day 30 after onset of diagnosis. Patients develop IgG and IgM isotypes of antibodies suggesting a key role for CD4+ T cells in isotype switching and memory responses after COVID-19 infection (30). It has been reported earlier that the SARS-CoV-2-specific antibodies along with SARS-CoV-2-specific helper and cytotoxic cells persist around 6-8 months (31). In the current study, we have analyzed adaptive T cell responses until day 28 after onset of diagnosis and showed the COVID antigen specific T cell responses and further time points can be evaluated in our future studies. Ten individuals who expired in this longitudinal study due to COVID-19 infection, all had COVID-19 specific antibody responses prior to death that is comparable to other patients, suggesting that their humoral and adaptive immune responses are inadequate to control and clear the infection. As we are observing higher responses at day 7 for AIM markers in deceased patients, it may due to more cytokine production (cytokine storm, CS), that is creating vulnerable immune responses in these patients. In literature, it has been shown that CS in COVID-19 infection is triggered by increased production of IL-6, IL-10, TNF-α, IFN-γ, etc (32). These can be produced by an uncontrolled immune response, like-continuous activation and expansion of immune cells, lymphocytes, and macrophages. Although cytokines can be produced by different immune cells like-DC, NK, B cells, along with T cells, CS could be a reason for obtaining high CD4^+^ T cells activation a few days before the participant’s death. There is evidence that SARS-CoV-2–specific T cells predominantly produced proinflammatory cytokines, like-effector and T helper 1 (Th1) cytokines, as well as Th2 and Th17 cytokines (33). We can assume that after CS some days were needed to develop severe systemic inflammation, other complications, and organ failure, which ultimately lead to death.

In summary, we found robust CD4^+^ AIM responses that were observed in COVID-19 patients and these responses are more in severe patients compared to asymptomatic and mild patients. Patients with moderate and severe disease developed higher levels of AIM responses that may help to generate good humoral responses to clear the infection in COVID-19 patients. In the patients who died it was high. Our data also suggests that T cell response plays role as function of disease severity and possibly recovery from Covid-19 infection and it can inform to develop or improve candidate vaccines to combat this novel coronavirus infection.

## Supporting information

Data Sheet

## Data Availability

The raw data supporting the conclusions of this article will be made available by the authors, without undue reservation

## Figure Legends

**Supplementary Figure 1:** Representative plot and gating strategy for T cell phenotyping antibody panel

**Supplementary Figure 2:** Representative plot and gating strategy for Mucosa Associated Invariant T (MAIT) cell antibody panel

**Supplementary Figure 3:** Representative plot and gating strategy for Activation Induced Marker (AIM) assay antibody panel

**Supplementary Figure 4:** Percentages of PBMC recovery of different disease groups after thawing at their day 1 (4A) and day 28 (4B). Number of PBMC (in million) per mL of blood from the disease groups at day 1 (4C) and day 28 (4D). Dunnett’s Multiple Comparison Tests were performed in both cases to compare between groups and *p<0.05.

**Supplementary Figure 5:** (A) Presentative plot for Fluorescence-activated cell sorting (FACS) gating for AIM+ (OX40+ CD40L+) cells gated on CD4+ T cell; (B) AIM+ CD4+ T cell reactivity in COVID-19 cases between the negative control (DMSO) and different antigen-specific stimulations (Spike, Non-spike MP, CMV, PHA). Wilcoxon matched-pairs signed rank test was performed to compare between groups. *p<0.05; **p<0.01; ***p< 0.001; ns: non-significant.

**Supplementary Figure 6:** (A) Presentative plot for Fluorescence-activated cell sorting (FACS) gating for AIM^+^ (CD69^+^ CD137+) cells gated on CD8^+^ T cell; (B) AIM^+^ CD8^+^ T cell reactivity in COVID-19 cases between the negative control (DMSO) and different antigen-specific stimulations (CD8-A MP, CD8-B MP, CMV, PHA). Wilcoxon matched-pairs signed rank test was performed to compare between groups. *p<0.05; **p<0.01; ***p<0.001; ns: non-significant.

**Supplementary Table 1:** Demographic information for participants of AIM assay

**Supplementary Table 2:** Antibody Panel for T cell phenotyping

**Supplementary Table 3:** Antibody Panel for MAIT cell phenotyping

**Supplementary Table 4:** Antibody Panel for NK cell phenotyping

**Supplementary Table 5:** Antibody Panel for Activation Induced Marker (AIM) assay

## ETHICS STATEMENT

The studies involving human participants were reviewed and approved by Ethical review committee of the International Centre for Diarrhoeal Disease Research, Bangladesh. The patients/participants provided their written informed consent to participate in this study.

## CONFLICTS OF INTEREST

A.S. is a consultant for Gritstone Bio, Flow Pharma, ImmunoScape, Avalia, Moderna, Fortress, Repertoire, erson Lehrman Group, RiverVest, MedaCorp, and Guggenheim. La Jolla Institute has filed for patent protection for various aspects of T cell epitope and vaccine design work. The authors declare that the research was conducted in the absence of any commercial or financial relationships that could be construed as a potential conflict of interest.

## AUTHOR CONTRIBUTIONS

FQ, TRB, FC, AA, TA designed, managed and supervised the study. AA, TA, IT, AH, MAS, AR, SG helped to collect the specimens. HAB, MHK, PCK, AH, SI performed the laboratory work and immunological analyses. TRB, HAB, MHK, PCK analyzed the data. FQ, TRB, DW, TS provided key reagents. TRB, HAB, MHK, PCK drafted the manuscript. FQ, TRB, SB, TS, JDC, DW, SB, TS, FC, AA, TA, IT reviewed and planned the manuscript. All authors contributed to the interpretation of results and critical review and revision of the manuscript and have approved the final version.

## ACKNOWLEDGEMENTS

This study was supported by Grant INV-018954 from the Bill and Melinda Gates Foundation, Global Emerging Leader Award (K43TW010362), R01 AI130378 of the National Institutes of Health (NIH), and icddr,b. This work has been supported by NIH contract 75N93019C00065 (A.S, D.W). The authors would like to thank Ministry of Health and Family Welfare (MOHFW) of Bangladesh, IEDCR, ideSHi, Kurmitola General Hospital and Mugda Medical College & Hospital for their continuous support. We also like to thank Daniela Weiskopf for providing the SARS-CoV-2 peptides for carrying out AIM assay. The authors would also like to express their sincere thanks to the staff members of icddr,b for their dedicated work in the field and laboratory during the pandemic situation. icddr,b is supported by the Governments of Bangladesh, Canada, Sweden, and the UK. The funders had no role in study design, data collection and analysis, decision to publish, or preparation of the manuscript.

## Notes

### Competing Interest Statement

The authors have declared no competing interest.

