## Supplementary material for "Upregulation of Activation Induced Cell Markers (AIM) among Severe COVID-19 patients in Bangladesh": Data Sheet

**Title:**

**Running title:** AIM assay in COVID-19 patients

[¶]Co-first author; [#]Senior author; [*]Corresponding author

*Dr. Firdausi Qadri, PhD
Senior Scientist and Head, Mucosal Immunology and Vaccinology Unit,
Infectious Diseases Division
International Centre for Diarrhoeal Disease Research, Bangladesh (icddr,b),
68, Shaheed Tajuddin Ahmed Sarani, Mohakhali, Dhaka 1212, Bangladesh.

**Supplementary Table 01: Demographic information for the participants of AIM assay**

| Variables | | COVID-19 patients (n=42) | | | | | Healthy Control (n=9) | Unexposed (Pre-pandemic) (n=10) |
|---|---|---|---|---|---|---|---|---|
| | | **Asymptomatic (n=5)** | **Mild (n=6)** | **Moderate (n=6)** | **Severe (n=15)** | **Expired (n=10)** | | |
| Mean Age (Years) | 44.7 (n=61) | 45 | 41.3 | 48.5 | 45.3 | 58.7 | 41.78 | 31.9 |
| Sex | Male (n=34) | 1 | 2 | 3 | 12 | 6 | 5 | 5 |
| | Female (n=27) | 4 | 4 | 3 | 3 | 4 | 4 | 5 |
| Blood Group | O+ (n=20) | 1 | 2 | 2 | 5 | 2 | 5 | 3 |
| | A+ (n=13) | 2 | 2 | 0 | 4 | 3 | 0 | 2 |
| | B+ (n=21) | 2 | 2 | 2 | 5 | 4 | 3 | 3 |
| | AB+ (n=6) | 0 | 0 | 2 | 1 | 1 | 0 | 2 |
| | O- (n=1) | 0 | 0 | 0 | 0 | 0 | 1 | 0 |

**Supplementary Table 02: Antibody Panel for T cell phenotyping**

| SL* | Marker | Fluorochrome | Clone | Company | Catalog# | Dilution |
|---|---|---|---|---|---|---|
| 1 | Live/Dead | (Fixable Near-IR) | - | Thermo Fisher | L10119 | 1:1000 [Step 1] |
| 2 | CD3 | Amcyan | SK7 | BD Biosciences | 339186 | 1:100 |
| 3 | CD19 | FITC | HIB19 | BD Biosciences | 555412 | 3:100 |
| 4 | CD4 | PerCP | SK3 | BD Biosciences | 347324 | 3:100 |
| 5 | CD8 | PECy7 | SK1 | BD Biosciences | 335787 | 1:100 |
| 6 | CXCR5 | BV421 | RF8B2 | BD Biosciences | 562747 | 2:100 |
| 7 | CD45RO | PE | UCHL1 | BD Biosciences | 555493 | 3:100 |
| 8 | CD27 | APC | O323 | Thermo Fisher | 17-0279-42 | 2:100 |

*SL = Serial Number

**Supplementary Table 03: Antibody Panel for MAIT cell phenotyping**

| SL* | Marker | Fluorochrome | Clone | Company | Catalog# | Dilution |
|---|---|---|---|---|---|---|
| 1 | Live/Dead | (Fixable Near-IR) | - | Thermo Fisher | L10119 | 1:1000 [Step 1] |
| 2 | CD3 | Amcyan | SK7 | BD Biosciences | 339186 | 1:100 |
| 3 | CD4 | PerCP | SK3 | BD Biosciences | 347324 | 3:100 |
| 4 | CD8 | PECy7 | SK1 | BD Biosciences | 335787 | 1:100 |
| 5 | TCR Vα7.2 | PE | 3C10 | Biolegend | 351706 | 2:100 |
| 6 | CD161 | APC | DX12 | BD Biosciences | 550968 | 2:100 |
| 7 | CD69 | PE-Cy5 | FN50 | BD Biosciences | 555532 | 2:100 |

*SL = Serial Number

**Supplementary Table 04: Antibody Panel for NK cell phenotyping**

| SL* | Marker | Fluorochrome | Clone | Company | Catalog# | Dilution |
|---|---|---|---|---|---|---|
| 1 | Live/Dead | (Fixable Near-IR) | - | Thermo Fisher | L10119 | 1:1000 [Step 1] |
| 2 | CD3 | PB | SP34-2 | BD Biosciences | 558124 | 1:100 |
| 3 | CD19 | APC Cy7 | SJ25C1 | BD Biosciences | 557791 | 1:100 |
| 4 | CD14 | APC Cy7 | MφP9 | BD Biosciences | 557831 | 1:100 |
| 5 | CD16 | FITC | 3G8 | BD Biosciences | 555406 | 2:100 |
| 6 | CD56 | PerCP Cy5.5 | B159 | BD Biosciences | 560842 | 2:100 |

*SL = Serial Number

**Supplementary Table 05: Antibody Panel for AIM assay**

| SL* | Marker | Fluorochrome | Clone | Company | Catalog# | Dilution |
|---|---|---|---|---|---|---|
| 1 | Live/Dead | (Fixable Near-IR) | - | Thermo Fisher | L10119 | 1:1000 [Step 1] |
| 2 | CD14 | BV785 | M5E2 | Biolegend | 301840 | 1:100 |
| 3 | CD16 | BV785 | 3G8 | Biolegend | 302046 | 1:100 |
| 4 | CXCR5 (CD185) | BV605 | J252D4 | Biolegend | 356930 | 1:100 |
| 5 | PD1 (CD279) | EF450 (PB) | MIH4 | ThermoFisher | 48-9969-42 | 2:100 |
| 6 | CD40L (C D154) | BV711 | 24-31 | Biolegend | 310838 | 2:100 |
| 7 | CD8 | PE-Cy7 | SK1 | BD Biosciences | 335787 | 1:100 |
| 8 | CD69 | PE-Cy5.5 | CH/4 | ThermoFisher | MHCD6918 | 2:100 |
| 9 | CD4 | Amcyan | SK3 | BD Biosciences | 339187 | 3:100 |
| 10 | CD137 | PE | 4B4-1 | Biolegend | 309804 | 1:100 |
| 11 | CD19 | FITC | HIB19 | BD Biosciences | 555412 | 1:100 |
| 12 | OX40 (CD134) | APC | Ber-ACT35 | Biolegend | 350008 | 1:100 |

*SL = Serial Number
