## Supplementary figures and images for "Upregulation of Activation Induced Cell Markers (AIM) among Severe COVID-19 patients in Bangladesh"

### Supplementary Figure 1

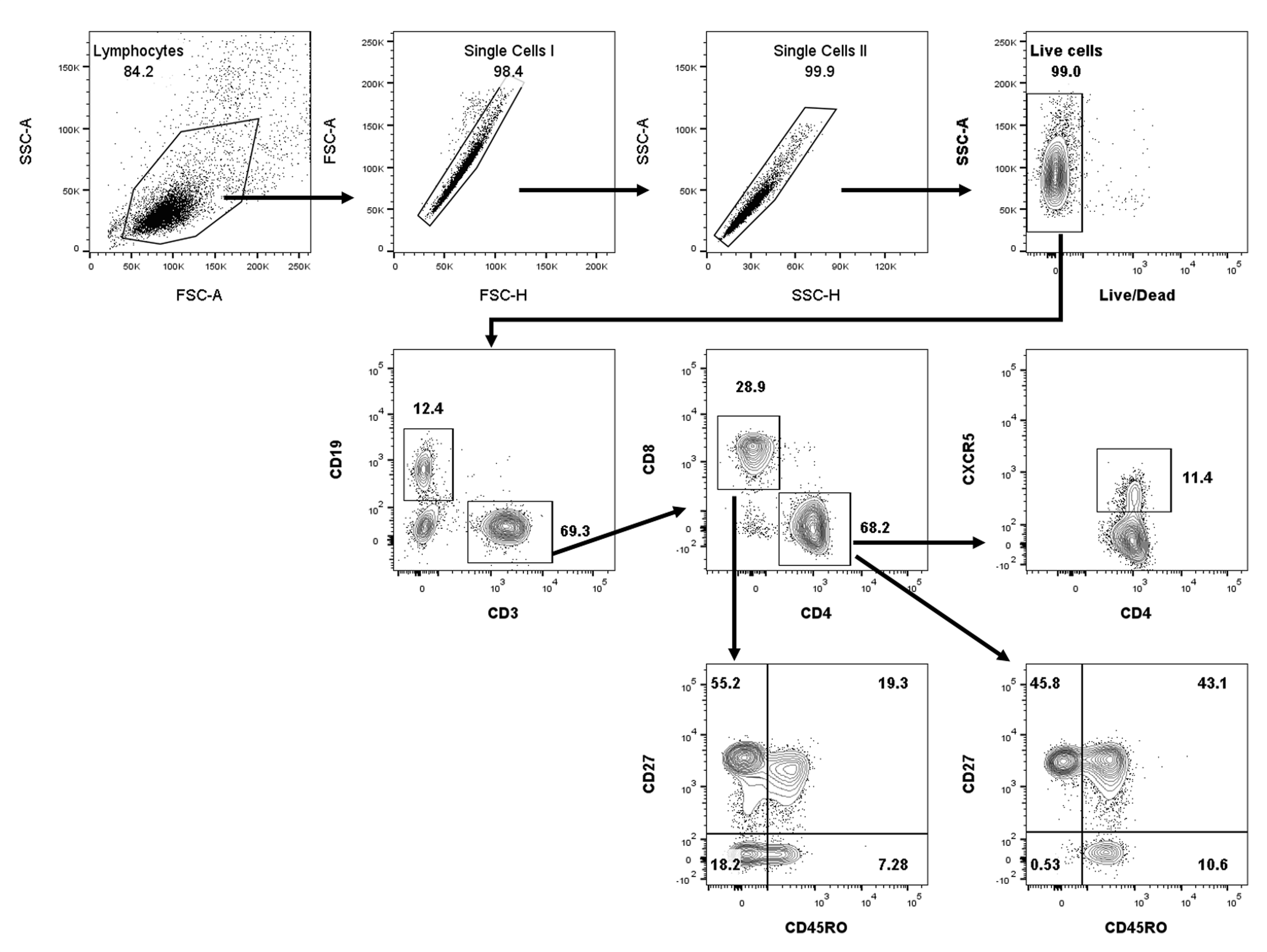

### Supplementary Figure 2

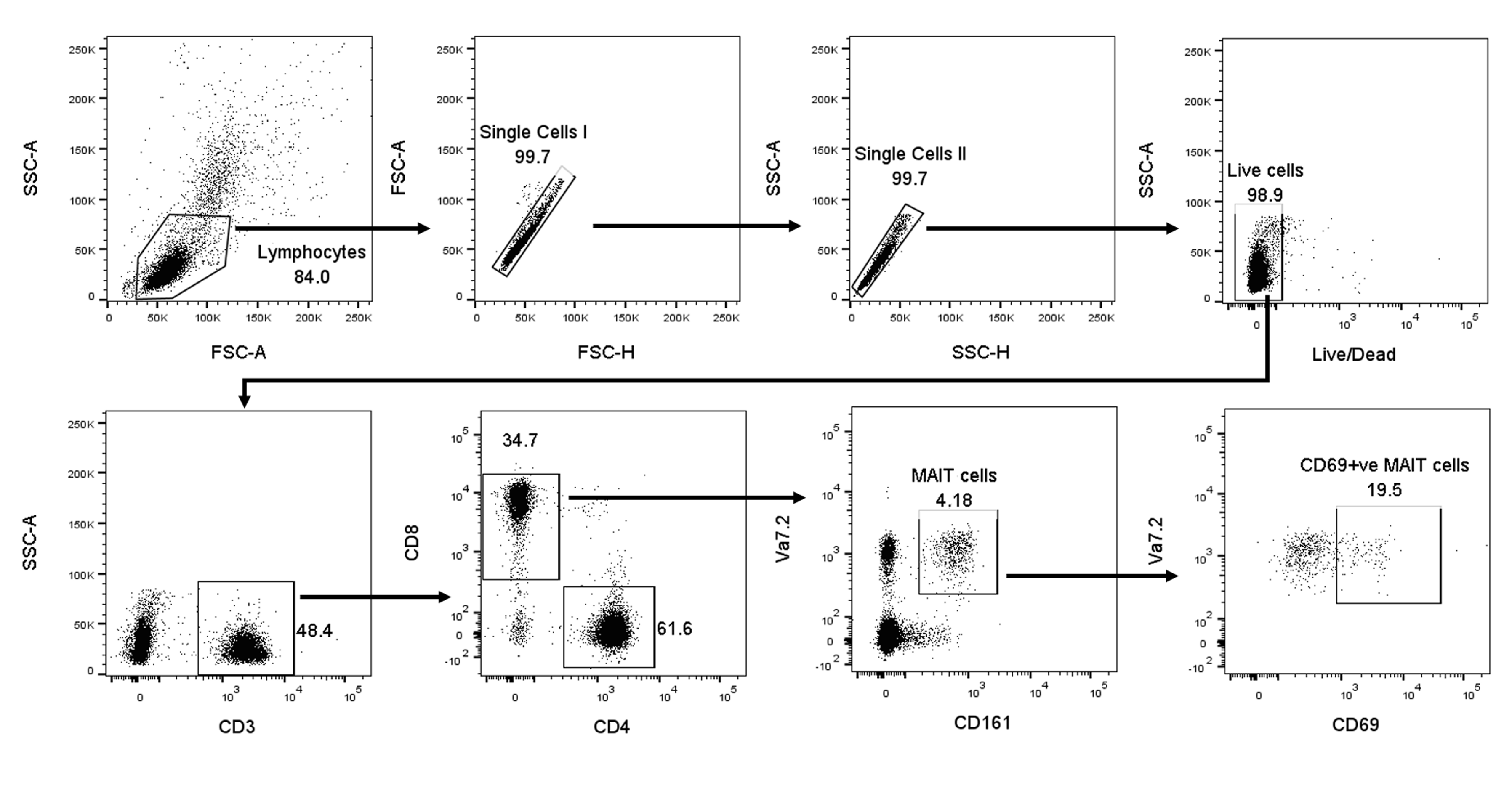

### Supplementary Figure 3

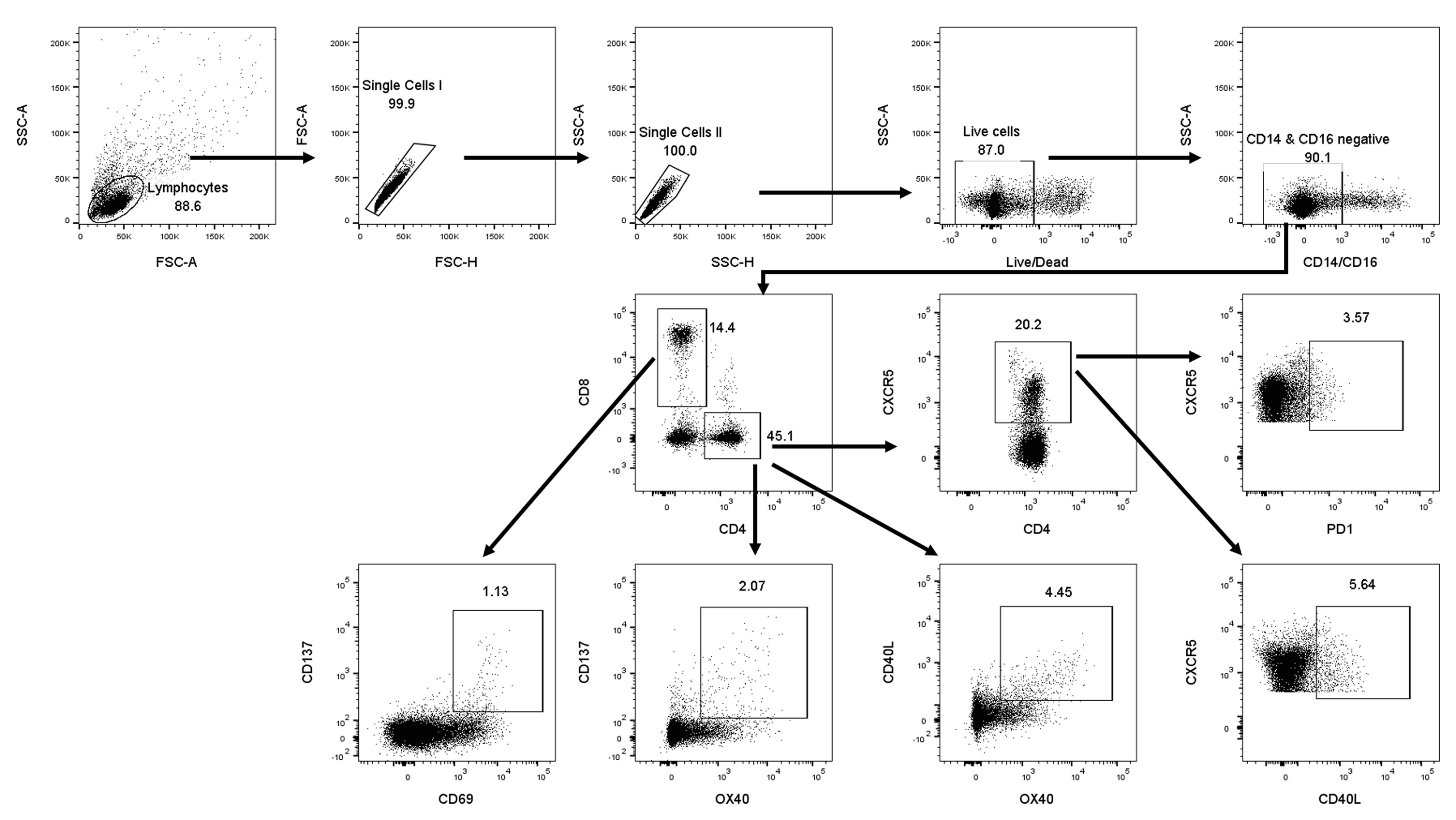

### Supplementary Figure 4

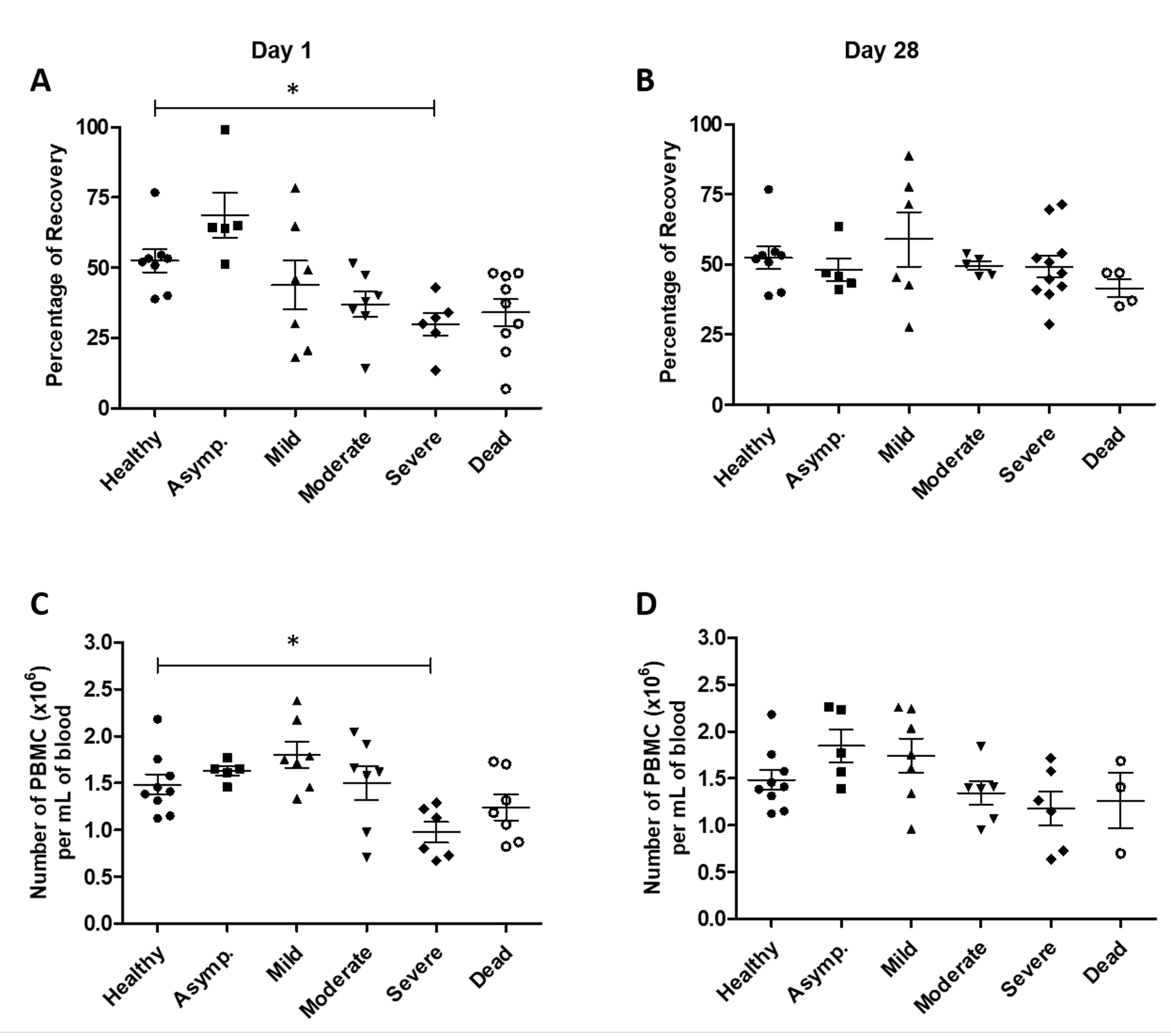

### Supplementary Figure 5

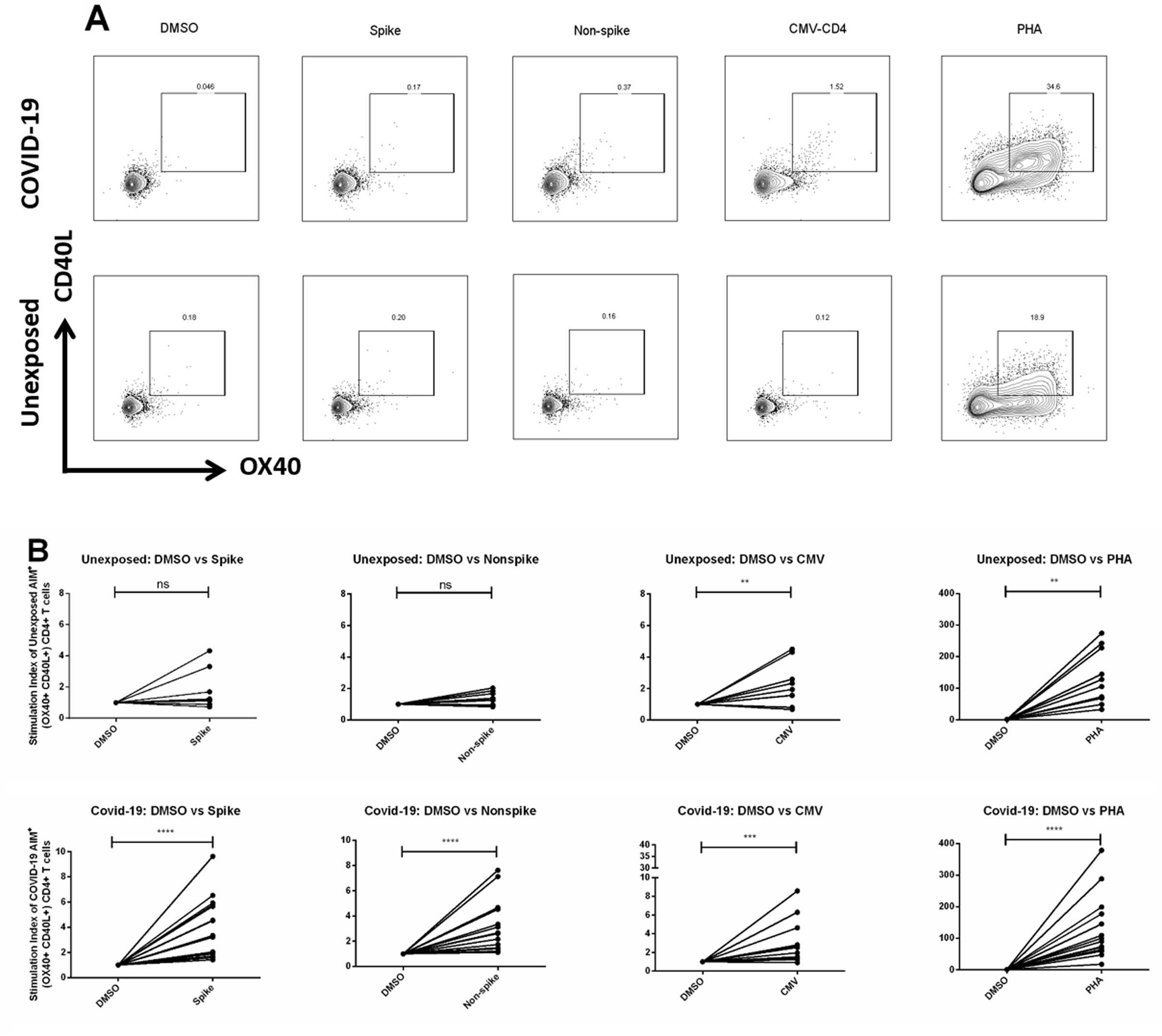

### Supplementary Figure 6

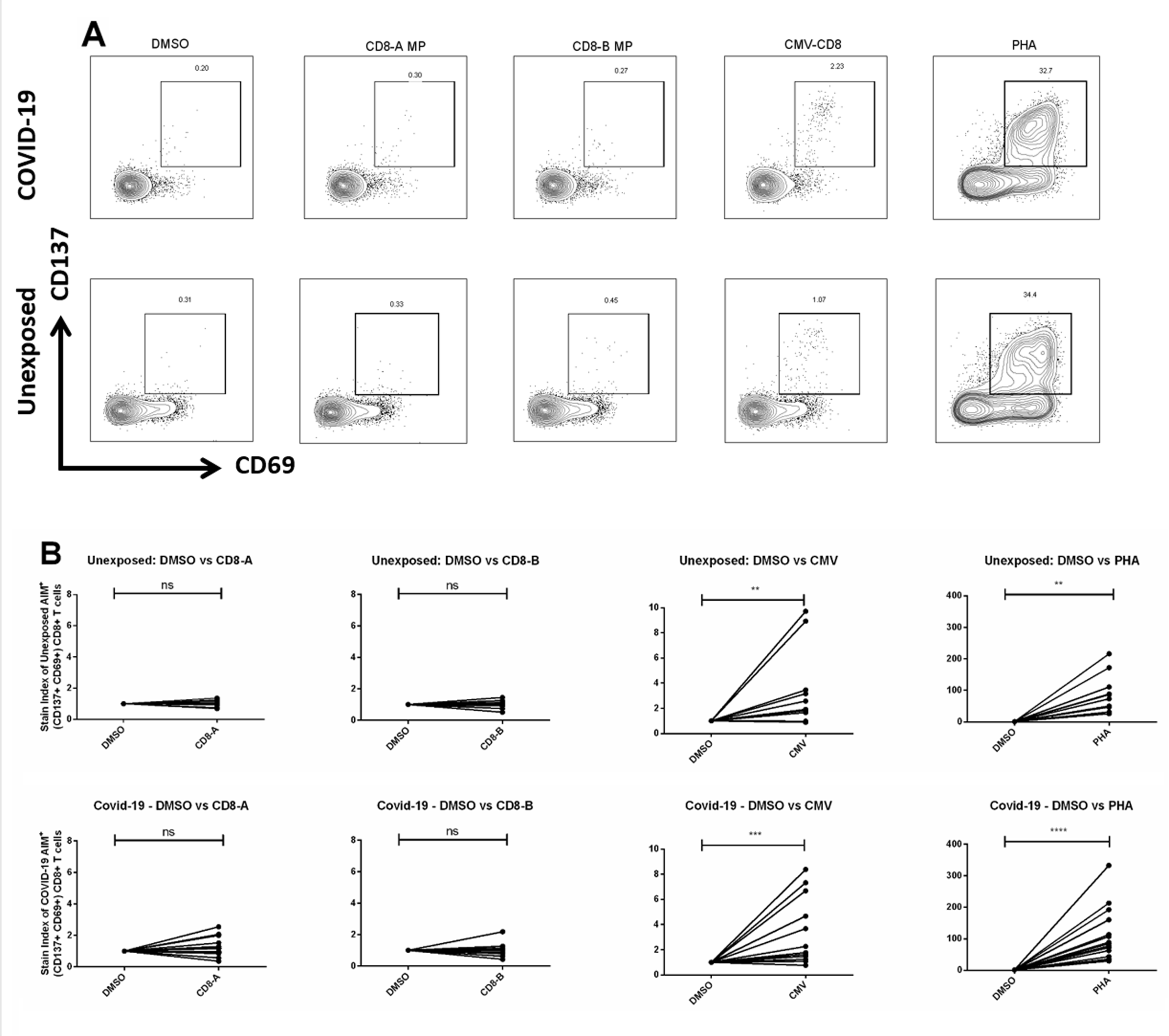
